# Genetically determined ancestry associates with morphological and molecular carotid plaque features

**DOI:** 10.1101/2025.06.07.25329125

**Authors:** Nima Fahim, Tim R. Sakkers, Floor B.H. van der Zalm, Joost Hoekstra, Dominique P.V. de Kleijn, Michal Mokry, Jose Verdezoto Mosquera, Gerard Pasterkamp, Hester M. den Ruijter, Clint L. Miller, Jessica van Setten, Sander W. van der Laan

## Abstract

Atherosclerosis, the main driver of cardiovascular disease (CVD), is influenced by a plethora of risk factors, including age, gender, and diabetes, that correlate with socio-economic status and may vary across ethnicities. These factors fail to fully explain observed ethnic disparities in CVD burden. For example, coronary artery calcification increases with age regardless of ethnicity, yet CAC is more prevalent in individuals of European descent. As these findings may be confounded by self-reported ethnicity, genome-informed ancestry offers a more accurate lens through which to study these ancestral differences. Yet, the biological basis of atherosclerotic plaque development and composition across ancestries remains essential underexplored.

We hypothesized that genetically determined ancestry is associated with morphological and molecular features of atherosclerotic plaques. Leveraging the Athero-Express Biobank Study, an ongoing Dutch cohort with deep histological and transcriptomic profiling of plaques, we analyzed data from 1,944 patients after genotype quality control and ancestry inference using principal component analysis against 1000 Genomes. Two ancestry groups were identified, European (n=1,866) and non-European (n=51), reflecting Netherlands’ migratory history.

Demographics were largely comparable between groups, however, ordinal logistic regression showed non-European ancestries had higher odds of increase plaque vulnerability (OR=1.67, 95% CI 1.01-2.77, p = 0.0450), a finding that remained robust after down sampling. Differential gene expression analysis highlighted *NLGN4X* and *CADM3* among the top differentially expressed genes, representing biologically relevant pathways related to synaptic and cell-cell adhesion. Pathway and single-cell enrichment analyses, including through integration with genome-wide association study data, further revealed consistent enrichment of inflammation-related biological processes and diseases.

Our findings support that genetic ancestry correlates with morphological and molecular plaque composition, with non-European patients showing more inflammatory, higher-risk plaque features, including inflammatory signatures. Increased ancestral diversity in vascular biology research is critical for understanding atherosclerotic pathophysiology and develop equitable and personalized therapeutic strategies.

## Letter

Cardiovascular disease (CVD) is the primary global cause of death, accounting for nearly 17.9 million deaths annually, with over 75% occurring in low- and middle-income countries^1^ . Atherosclerosis, its main driver, is influenced by gender, aging, smoking, hypertension, family history and genetics, type 2 diabetes, lifestyle, and dyslipidemia – risk factors that correlate with socio-economic status and may vary across ethnicities^2–4^. However, they fail to fully explain observed ethnic disparities in relative rates of CVD^3,4^. For example, coronary artery calcification (CAC) – a marker of subclinical atherosclerosis – steadily increases with age, irrespective of gender or ethnicity, yet CAC is more prevalent in individuals of European descent, and more so among men compared to women^3–6^. These results may be confounded by self-reported ethnicity, however, genome-informed ancestry has been shown to correlate with subclinical atherosclerosis^5^. Thus, investigating the biological basis of atherosclerotic plaque development across ancestries remains essential.

Histological analysis is currently the gold standard method to evaluate atherosclerotic plaque composition^7,8^. High-risk lesions are characterized by variable calcific, inflammatory, lipid, and thrombotic components increasing risk of rupture and ischemic events^7,9^. While pathological studies revealed a higher extent of calcified coronary lesions in individuals of European ancestry^8^, it remains unclear whether this extends to other morphological^9^ or molecular plaque features. We hypothesized that genetically determined ancestry is associated with plaque composition, independent of risk factors.

We leveraged the Athero-Express (AE) biobank study^7^ – a Dutch cohort with deep histological and molecular profiling^10–12^ of plaques. After quality control and ancestry inference using principal component analysis (PCA, relative to the 1000G phase 3 reference, b38), data for 1,944 patients remained, forming two groups: European (n=1,866) and non-European (n=51), reflecting Netherlands’ migratory history^13–15^ (Error! Reference source not found., **panel 1**). Demographics, including age, gender, diabetes prevalence, hypertension, BMI, renal function, cholesterol and hsCRP levels, symptom prevalence and smoking behavior, did not significantly (p > 0.05, ***Figure 1*, panel 2**) differ between Europeans and non-Europeans.

**Figure 1:**
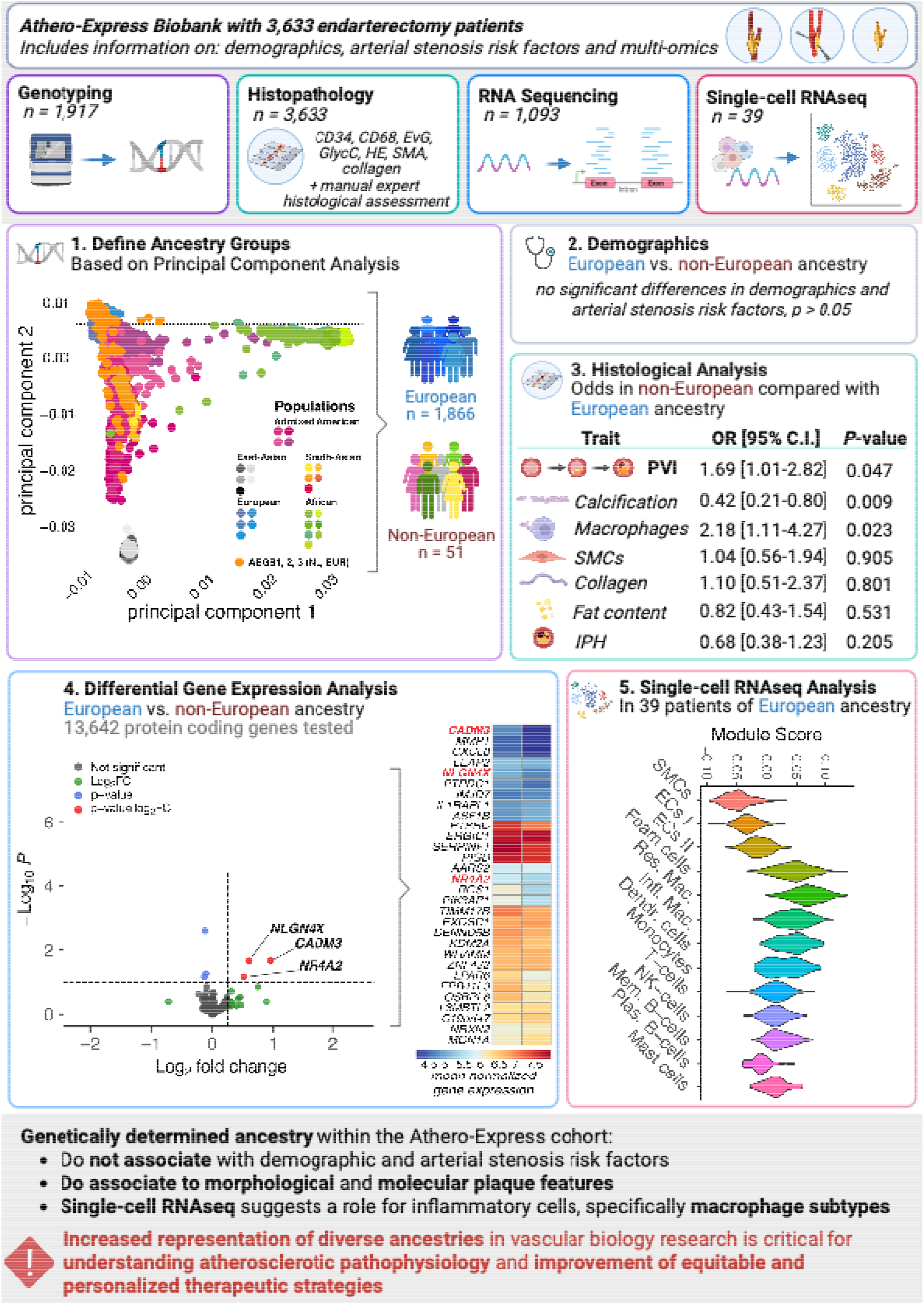
Genetically determined ancestry is associated with morphological and molecular plaque features. The Athero-Express Biobank Study (www.atheroexpress.nl) includes atherosclerotic plaque samples from 3,633 patients undergoing endarterectomy^7^. Routinely plaques are paraffin embedded, decalcified, and histologically and immunohistochemically stained for endothelial cells (CD34), macrophages (CD68), elastin (Elastic von Giesen, EvG), red blood cell content (Glycophorin C, GlycC), overall structural morphology (Hematoxylin and Eosin, HE), smooth muscle cells (SMA), and collagen (picrosirius red, SR)^7^. A subset of patients (n=1,917) were genotyped in three consecutive experiments using commercial genotyping platforms^10^; community standard quality control as applied^26^ and missing genotypes were imputed against the TOPMed reference (b38)^27^. Each experiment was named consecutively: Athero-Express Genomics Study 1, 2, and 3 (AEGS1, AEGS2, AEGS3). From 1,093 plaque samples RNA was isolated in two consecutive experiments, overlapping both histological and genotyped data (n=1,028), and a Celseq2 experimental protocol was adapted for bulk-tissue RNA sequencing described earlier^11^. **(1) Genetic principal component analysis (PCA)** against the 1000G phase 3 (version 5, b38) reference populations. Each population is grouped in a superpopulation: *Admixed American* (purple-pink-tinted bullets) including Mexican Ancestry from Los Angeles USA, Puerto Rican from Puerto Rica, Colombian from Medellian, Colombia, Peruvian from Lima, Peru; *East-Asian* (gray-tinted bullets) including Han Chinese in Bejing, China (CHB), Japanese in Tokyo, Japan (JPT), Southern Han Chinese, China (CHS), Chinese Dai in Xishuanagbanna, China (CDX), and Kinh in Ho Chi Minh City, Vietnam (KVH); *South-Asian* (red-yellow-tinted bullets) including Gujarati Indian from Houston, Texas, Punjabi from Lahore, Pakistan, Bengali from Bangladesh, Sri Lankan Tamil from the UK, and Indian Telugu from the UK; *European* (blue-tinted bullets) including Utah Residents (CEPH) with Northern and Western European ancestry, Toscani in Italia, Finnish in Finland, British in England and Scotland, and Iberian population in Spain; and *African* (green-tinted bullets) including Yoruba in Ibadan, Nigera, Luhya in Webuye, Kenya, Mandinka in The Gambia, Mende in Sierra Leone, Esan in Nigera, American’s of African Ancestry in SW USA, and African Carribean in Barbados. The orange-tinted bullets represent the AEGS1, AEGS2 and AEGS3 patients. The PCA revealed two distinct groups, 1,866 patients from European ancestry, and 51 patients from non-European ancestry. **(2) Demographic analyses** revealed no significant differences on risk factors (see main text) between these groups. **(3) Histological analyses** using ordinal and logistics regression, both univariate and multivariate (see text) showed a significant correlated with the plaque vulnerability index (PVI) with non-Europeans having higher odds of increases PVI, i.e. a more unfavorable plaque composition (comprising more intraplaque hemorrhage [IPH], inflammation, more fat content, and fewer collagen, and smooth muscle cells [SMCs]). Sensitivity analyses of individual morphological features, including calcification, macrophage and SMCs, collagen and fat content, and IPH revealed lower odds of calcified plaques and higher odds of inflammatory plaques in non-Europeans. **(4) The differential gene expression analysis** in the samples with both genetic and RNAseq data (n=1,028), utilizing only protein-coding genes mapped against the GRCh38 (hg38) genome assembly with DESeq2 (baseMean > 10), revealed three genes, *NLGN4X, NR4A2*, and *CADM3*, significantly differentially expressed when comparing Europeans (reference) with non-Europeans (volcano plot, left, see also text). The heatmap shows the top 30 differentially expressed genes (see text), where *NLGN4X, NR4A2*, and *CADM3* are significantly upregulated. Note: For consistency only the results of the multivariate analyses in (3) and (4), corrected for age, gender, genotype platform, and year of surgery, are displayed. **(5) Single-cell RNAseq based module score analysis** of the differentially expressed genes in 39 samples of European ancestry (non-overlapping with the aforementioned 1,028 samples, described elsewhere^25^, and a subset is accessible at www.plaqview.com^28^) support the notion of the involvement of inflammatory cells, specifically foam cells, resident macrophages (*Res. Mac*.), and inflammatory macrophages (*Infl. Mac*.), but not T-cells, natural killer cells (NK-cells), memory B-cells (*Mem. B-cells*) or plasma B-cells (*Plas. B-cells*). Created in BioRender. Van der laan, S.W. (2025) https://BioRender.com/yat6tjd.

To assess plaque vulnerability, we used the plaque vulnerability index (PVI, a histological score incorporating collagen, macrophages, smooth muscle cells, and lipid content)^7^. Each trait was scored as stable or unstable, yielding a cumulative vulnerability score from 0 to 4. Those with non-European ancestries had higher odds of increased PVI (OR=1.67, 95% CI 1.01-2.77, p = 0.045, ***Figure 1*, panel 3**), even after adjusting for age, gender, year of surgery, and genotyping platform in ordinal logistic regression (OR=1.69, 95% CI 1.01-2.82, p = 0.047). Sensitivity analyses using down-sampling (20-fold) of Europeans revealed a median OR=1.13, 95% CI 1.03-1.25, 18/20, p_binomial_ < 0.001) indicating our results are robust with regards to the association with PVI. Further dissection of the role of individual plaque composition features revealed that non-Europeans showed lower odds of calcification (OR=0.42, 95% CI 0.21-0.80, p = 0.009) and higher odds of macrophage-rich plaques (OR=2.18, 95% CI 1.11-4.26, p = 0.023). Which was especially true for male gender: OR=0.75 95% CI 0.61-0.93 p=0.009 for calcification, and 1.4 95% CI 1.41-2.15 for macrophages, p=1.9×10^-7^, respectively. This is consistent with earlier work that men of European descent have increased risk for CAC, underscoring that our study is sufficiently powered to detect established gender-related differences in plaque composition^6^. Our findings suggest a potential ancestry-related difference in inflammatory versus calcified plaque phenotypes, where non-European patients exhibit higher inflammation (macrophages) and lower mineralization (calcification), consistent with the elevated overall plaque vulnerability observed.

Transcriptomic analyses of 1,028 plaques revealed 1,146 genes differentially expressed (p_nominal_<0.05) out of 13,642 (p_binomial_ = 1.11×10^-62^) between ancestry groups, including *NLGN4X* and *CADM3* (log_2_FC=+0.61 and +0.94, respectively, FDR < 0.09) linked to synaptic and cell-cell adhesion^16^. Among the top ancestry-associated transcripts, *MON1A, NRXN2, WHAMM*, and several mitochondrial and Golgi-associated genes showed higher expression in Europeans. In contrast, non-European plaques exhibited higher expression of neuronal adhesion, and immune- and inflammation-related genes including *MMP1, NR4A2, CADM3*, and *NLGN4X* (Error! Reference source not found., **panel 4**). To further investigate the underlying mechanisms, we performed gene-set enrichment analyses using fgsea^17^ and MSigDB^18^ hallmark pathways revealing inflammation-related signatures including epithelial-to-mesenchymal transition, inflammatory response, apoptosis, and coagulation (p_adjusted_ < 0.05). To contextualize the ancestry-associated transcriptional changes we used Enricher^19^ to assess enrichment of gene-disease associations (DisGeNET^20^) showing overrepresentation of genes affecting diseases where the immune-driven diseases, such as inflammatory bowel disease, ulcerative colitis, neoplasms, arthritis, and abdominal aortic aneurysms (p_adjusted_ < 0.05). Assessing the enrichment of the DEGs in plaque-derived single-cell RNAseq (n=39, only available of Europeans) data suggest a role for inflammatory cells, specifically macrophages (***Figure 1***, panel 5). Enrichment against GWAS Catalog^21^ further highlighted associations with circulating myeloid cell traits such as neutrophil-to-lymphocyte ratio, eosinophil percentage and mean corpuscular hemoglobin (p_adjusted_ < 0.05). To assess cardiovascular relevance, we tested for enrichment of ancestry-associated genes against those linked to coronary artery disease^22^, CAC^23^, and carotid IMT^24^ using MAGMA based gene-level GWAS summary statistics^25^. All traits showed higher-than-expected overlap via binomial testing (CAC: 102 nominally significant in CAC GWAS/1,074 nominally significant in our DGEA; CAD: 145/1,077; cIMT: 98/1,077; all p < 2.3×10^−1^0). These complimentary enrichment analyses underscore the robustness of our findings and suggest biological relevance of the ancestry-associated transcriptional difference to atherosclerotic disease.

Despite the strengths of this study – including histological analyses, genetic inference and transcriptomic data – several limitations should be noted. While this is the largest ancestry-focused multimodal study of carotid plaques to date, the small non-European sample limits generalizability. The cohort reflects advanced disease, limiting the extrapolation to earlier subclinical stages of atherosclerosis. Socio-economic status was not directly assessed and may confound ancestry-related differences, although demographics did not differ between ancestry groups.

Our findings support that genetic ancestry is associated with plaque composition with non-European patients showing higher-risk plaque features. Morphological analyses point to increased inflammation and reduced mineralization in non-Europeans, which is supported by transcriptomic data showing broad inflammatory and immune-related transcriptional shifts. Increased representation of diverse ancestries in vascular biology research is critical for understanding atherosclerotic pathophysiology and to improve the design of equitable and personalized therapeutic strategies.

## Data Availability

All data used in the present study are available through a DataverseNL repository. The code for this work are available in a GitHub repository.

https://doi.org/10.34894/4IKE3T

https://github.com/CirculatoryHealth/PlaqueMorphology_Ancestry_Public

## CREDiT authorship contributions

This is based on the PLoS Genetics format: https://journals.plos.org/plosgenetics/s/authorship#loc-author-contributions

## Acknowledgements

We are thankful for the support of the Leducq Fondation ‘PlaqOmics’ (18CDV-02) and ‘AtheroGen’, and the Chan Zuckerberg Initiative ‘MetaPlaq’. The research for this contribution was made possible by the AI for Health working group of the EWUU alliance (https://aiforhealth.ewuu.nl/). The collaborative project ‘Getting the Perfect Image’ was co-financed through use of PPP Allowance awarded by Health∼Holland, Top Sector Life Sciences & Health, to stimulate public-private partnerships.

## Disclosures

Dr. Sander W. van der Laan and Gerard Pasterkamp received Roche funding for unrelated work. Roche had no part in this study, neither in the conception, design, and execution of this study, nor in the preparation and contents of this manuscript. Dr. Clint L. Miller received grant support from AstraZeneca for work unrelated to the current study.

ChatGPT for macOS was used to check the text for spelling, grammar and syntax; after use we edited the texts and the authors take full responsibility for the content of this manuscript. Adobe Illustrator 2025 v29.7 (Adobe Inc., San Jose, CA, USA) was used to improve legibility of the fonts in graphs.

## Code and data

The data and used for these analyses are available through a Dataverse repository (https://doi.org/10.34894/4IKE3T), and a GitHub repository (https://github.com/CirculatoryHealth/PlaqueMorphology_Ancestry_Public).

